# Far from MCAR: obtaining population-level estimates of HIV viral suppression

**DOI:** 10.1101/19012781

**Authors:** Laura B. Balzer, James Ayieko, Dalsone Kwarisiima, Gabriel Chamie, Edwin D. Charlebois, Joshua Schwab, Mark J. van der Laan, Moses R. Kamya, Diane V. Havlir, Maya L. Petersen

## Abstract

**Background:** Population-level estimates of disease prevalence and control are needed to assess the effectiveness of prevention and treatment strategies. However, available data are often subject to differential missingness. Consider population-level HIV viral suppression: proportion of all HIV-positive persons who are suppressing viral replication. Individuals with measured HIV status, and, among HIV-positive individuals, those with measured viral suppression are likely to differ from those without such measurements.

**Methods:** We discuss three sets of assumptions sufficient to identify population-level suppression over time in the intervention arm of the SEARCH Study (NCT01864603), a community randomized trial in rural Kenya and Uganda (2013-2017). Using data on nearly 100,000 participants, we compare estimates from an unadjusted approach assuming data are missing-completely-at-random (MCAR); stratification on age group, sex, and community; and, targeted maximum likelihood estimation (TMLE) with Super Learner to adjust for baseline and time-updated predictors of measurement.

**Results:** Despite high annual coverage of testing, estimates of population-level viral suppression varied by identification assumption. Unadjusted estimates were most optimistic: 50% of HIV-positive persons suppressed at baseline, 80% at Year 1, 85% at Year 2, and 85% at Year 3. Stratification on baseline predictors yielded slightly lower estimates, and full adjustment reduced estimates further: 42% of HIV-positive persons suppressed at baseline, 71% at Year 1, 76% at Year 2, and 79% at Year 3.

**Conclusions:** Estimation of population-level disease burden and treatment coverage require appropriate adjustment for missingness. Even in “Big Data” settings, estimates relying on the MCAR assumption or baseline stratification should be interpreted with caution.

## INTRODUCTION

Accurate population-level estimates of disease prevalence and treatment coverage are needed to quantify disease burden and evaluate the success of programs for epidemic control. The data available to inform such estimates, however, are often susceptible to differential measurement. In other words, the missing-completely-at-random (MCAR) assumption rarely, if ever, holds.^1–4^ The field of HIV provides an illustrative example. Consider the UNAIDS 90-90-90 target: 90% of all HIV-positive persons should know their status; 90% of those who know their status should be receiving antiretroviral therapy (ART); and 90% of those receiving ART should have suppressed HIV viral replication.^5^ Multiplying these proportions together yields an overall target, referred to here as “population-level suppression” - 73% of all HIV-positive persons should be suppressing HIV viral replication (Appendix). This target reflects the HIV care “cascade” from diagnosis, through treatment initiation and retention, to viral suppression.

While population-level suppression is widely used in assessing HIV care strategies, two recent systematic reviews noted the variability in both data quality and statistical approaches used for assessment.^6,7^ In particular, Granich *et al*. remarked on the challenges posed by incomplete data and inconsistent methodology, while Sabapathy *et al*. proposed a template to standardize data collection and evaluation. In this manuscript, we provide an in-depth demonstration of the methods used to estimate population-level suppression in the SEARCH Study, a cluster randomized trial in rural Kenya and Uganda (NCT01864603).^8,9^ We approach the missing data problem with a causal framework to define target parameters with counterfactuals, state identifiability assumptions sufficient to translate these targets into statistical quantities, and estimate the resulting statistical parameters.^1–4,10–14^ We refer the reader to companion papers for details on the trial.^8,9^

## METHODS

In general, the total number of HIV-positive persons in a population is unknown, and individuals for whom HIV status is known are not necessarily representative of the general population. If testing (e.g. health-seeking behavior) is related to HIV status, unadjusted estimates of prevalence (i.e. the proportion of those with HIV among those with known status) are likely to be biased, even in the context of community-wide testing, as was implemented in recent Universal-Test-and-Treat trials.^9,15–18^

Likewise, measurement of plasma HIV RNA levels (viral loads) among HIV-positive individuals is generally incomplete and often depends on factors associated with viral suppression status. For example, if viral loads are only measured at HIV clinic visits, then viral suppression among individuals with known status will overestimate suppression among all HIV-positive persons, including newly diagnosed individuals who have not yet linked to care and previously diagnosed individuals who have never linked or have dropped-out of care. These familiar missing data challenges can be illustrated with a directed acyclic graph or another causal modeling approach (Figure 1).^14,19–24^

**Figure 1:**
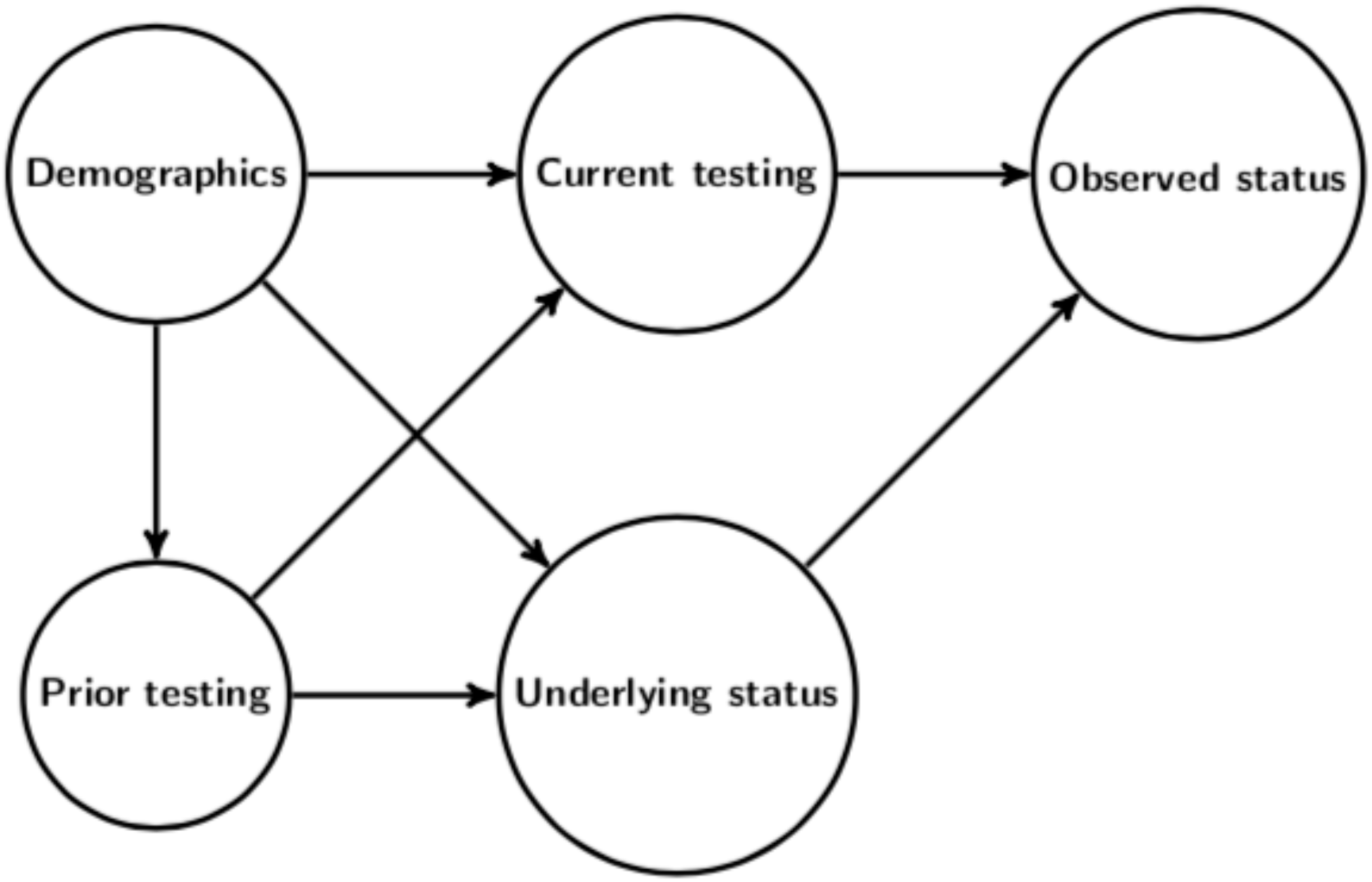
Simplified directed acyclic graph to represent the challenges posed by incomplete HIV testing. Demographics and prior testing are common causes of current testing (the hypothetical intervention node) and underlying HIV status (possible unobserved), both of which impact observed HIV status. Analogous challenges arise due to incomplete measurement of suppression among HIV-positive persons.

Overcoming these challenges requires knowledge of the data generating process. Consider the 16 communities in the intervention arm of the SEARCH Study. After a door-to-door census, community-wide testing was conducted annually through multidisease health fairs, followed by out-of-facility testing for residents who did not attend the fair.^25,26^ Participants were linked over successive years with a fingerprint biometric. Prior diagnosis of HIV and ART use were ascertained through linkage to clinic records.^8,27^ A re-census was conducted three years after follow-up to determine interim deaths, out-migrations, and in-migrations.^9^

With this measurement scheme in mind, we describe the methods used in Petersen *et al*.^8^ and Havlir *et al*.^9^ to characterize viral suppression in the intervention arm at the time of community-wide testing: study baseline *t=0*, and annually thereafter *t=*{*1,,3*}. These cross-sectional analyses provide snapshots of population-level suppression among an open cohort of adult (≥15years) residents (allowing for entry due to age and in-migration, and exit due to death or outmigration). We note estimating viral suppression among a closed cohort of baseline HIV-positive residents is a distinct goal, resulting in a different causal parameter, identifiability assumption, and estimation approach.^8,9^

### Causal parameters

Let 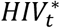 be an indicator that an individual is HIV-positive at time *t*, irrespective of whether serostatus is measured. Likewise, let 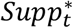 be a possibly unmeasured indicator of HIV viral suppression (<500cps/mL) at time *t*. Population-level suppression is the conditional probability of viral suppression given HIV-positive status: 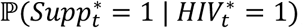, or equivalently, the joint probability of being HIV-positive with suppression, divided by the probability of being HIV-positive (i.e. HIV prevalence):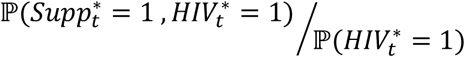.

Ideally, anyone not already known to be HIV-positive (i.e. previously HIV-negative or HIV-unknown) would be tested at time *t*. Of course, this is never the case; further, missingness inherently depends on underlying HIV status - the status of an HIV-negative individual who does not test at *t* is unknown, whereas the status of an HIV-positive individual not seen at *t* might be known from prior testing. The problem is intensified after multiple rounds of community-wide testing, which provide multiple opportunities for prevalent HIV-positive persons to be diagnosed. To avoid this inherent dependence, we define *TstHIV*_*t*_ as an indicator that an individual was seen at community-wide testing and had “known” HIV status at time *t* - due to a negative test result at time *t*, or a positive result at or before time *t*. We define observed HIV status as 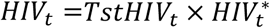.

As with HIV testing, viral load measurement is incomplete; HIV-positive persons on whom a viral load is measured may differ systematically from HIV-positive persons who are missing a viral load. Define *TstVL*_*t*_ as an indicator of viral load measurement at time *t*, and define observed viral suppression as 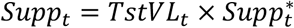.

### Three sets of identifiability assumptions

In the above, population-level suppression was expressed in terms of underlying indicators of HIV seropositivity and viral suppression: 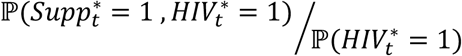.We now present three sets of identifiability assumptions to write the numerator and denominator in this expression as parameters of the observed data distribution.

#### Unadjusted

Suppose we are willing to assume that HIV prevalence among those seen at time *t* is representative of HIV prevalence among those not seen, and that viral suppression among HIV-positive persons with viral load measurement at time *t* is representative of suppression among HIV-positive persons without viral load measurement at that time. More formally, we are making the following randomization assumptions as applied to missing data: 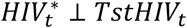 and 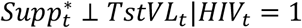 If these assumptions hold, the numerator of population-level suppression is identified as

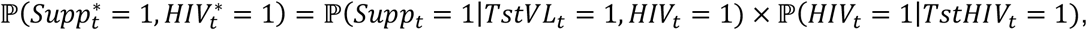

and denominator as 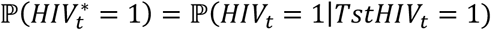.^27^ Taking the ratio of these yields the unadjusted statistical parameter:

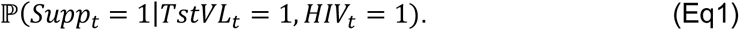

#### Baseline adjustment

We can weaken the above assumptions by conditioning on baseline covariates. Specifically, let *B* denote mutually exclusive and exhaustive strata defined by age group, sex, and community of residence. Now suppose within each strata *b*, HIV prevalence among those seen at *t* is representative of prevalence among those not seen, and within each strata *b*, suppression among HIV-positive persons with viral loads measured at *t* is representative of suppression among HIV-positive persons without measured viral loads. More formally, we assume 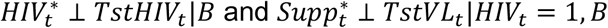.

Under these assumptions on missingness, we obtain the G-computation identifiability result,^28^ corresponding to a hypothetical, dynamic intervention to first ensure knowledge of HIV status and then to ensure measurement of viral loads among HIV-positive persons.^29–31^ Specifically, the proportion of the population that is HIV-positive and suppressed is identified as

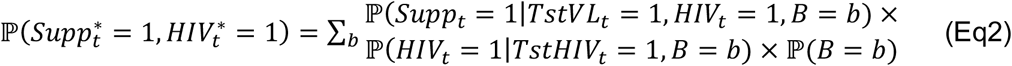

In words, this is the strata-specific probability of viral suppression, given measurement and HIV-positive status; multiplied by the strata-specific probability of being HIV-positive, given measurement; and then standardized with respect to the distribution of strata. Identification of the denominator, population-level prevalence, follows from the above:

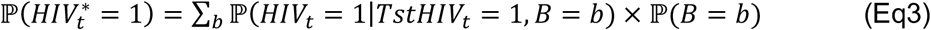

By taking the ratio of the numerator (Eq2) to the denominator (Eq3), we obtain a baseline-adjusted statistical parameter corresponding to population-level suppression under the above assumptions.

For the conditioning sets to be well-defined, we also require the positivity assumption.^11,32^ Irrespective of age, sex, and community, there must be a positive probability of being seen with known HIV status 𝕡(*TstHIV*_*t*_ = 1 |*B*) > 0, and for every strata in which some proportion of HIV-positive persons are seen, there must be a positive probability of viral load measurement, regardless of the stratification factors: 𝕡(*TstVL*_*t*_ = 1 |*HIV*_*t*_ = 1, *B*) > 0.

#### Time-varying adjustment

While stratifying on certain baseline characteristics weakens our assumptions on missingness, there may be many other variables potentially impacting the probability of being tested, underlying HIV status, and viral suppression among HIV-positive persons. In particular, ART use is a key determinant of viral suppression and may also be predictive of viral load measurement.

Define *ART*_*t*_ as an indicator of ART initiation prior to time *t*, and let *X*_*t*_ denote the remaining observed variables that are potentially predictive of both viral suppression and measurement: the full set of baseline demographics (e.g. age, sex, marital status, education, occupation, alcohol use, mobility, wealth index, and community) and prior HIV testing and suppression. While viral suppression without ART is possible, the UNAIDS target is focused on ART-induced suppression.^5^ Therefore, we set 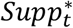 to zero for persons not on ART - acknowledging that incomplete capture of ART use will lead to underestimation of suppression. For HIV-positive persons who have initiated ART, we assume that conditional on the baseline and time-updated covariates *X*_*t*_, suppression among those with a viral load measured during annual testing is representative of suppression among those with a missing viral load. More formally, we assume 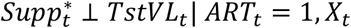.

We also require the positivity assumption; all HIV-positive individuals who have initiated ART have a positive probability of having their viral load measured, regardless of their baseline and time-updated covariates: 𝕡(*TstVL*_*t*_ = 1 |*ART*_*t*_ = 1, *X*_*t*_) > 0 a.e.. Under these assumptions, we have the G-computation identifiability result corresponding to a hypothetical intervention to ensure viral load measurement among ART initiators:^28^

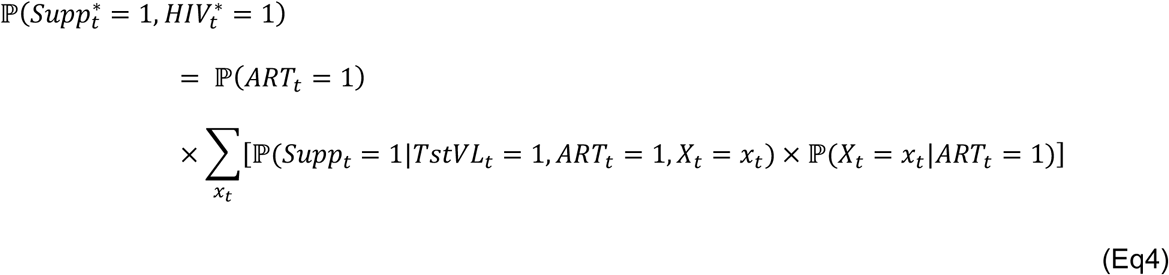

where the summation generalizes to an integral for continuous covariates. In words, this is the proportion of individuals who have started ART (and are, by implication, HIV-positive) in the total population (including both HIV-positive and HIV-negative persons) multiplied by the adjusted probability of being suppressed and measured, given prior ART initiation.

For the denominator of HIV prevalence, we also consider an expanded adjustment set *L*_*t*_, consisting of all baseline demographics and prior HIV testing (e.g. number and location). For the subgroup without a prior HIV diagnosis, we assume that conditional on *L*_*t*_, HIV prevalence among those tested at *t* is representative of HIV prevalence among those not tested, or more formally, 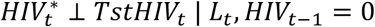. We further assume positivity; previously undiagnosed persons have some chance of being tested regardless of their *L*_*t*_ values: ℙ(*TstHIV*_*t*_ = 1 |*HIV*_*t* − 1_ = 0, *L*_*t*_) > 0 a.e.. Under these assumptions, we have the G-computation identifiability result corresponding to a hypothetical intervention to ensure HIV status is known:^28^

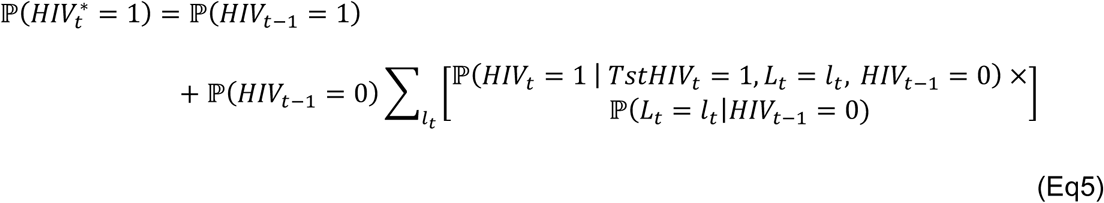

where the summation generalizes to an integral for continuous covariates. In words, this is the proportion of the population previously known to be HIV-positive plus the adjusted proportion of the population newly known to be HIV-positive.

Taking the ratio of the numerator (Eq4) to the denominator (Eq5) yields a fully-adjusted statistical parameter for population-level suppression under the above assumptions.

### Estimation approaches

The unadjusted parameter (Eq1) can be estimated with the empirical proportion of the population with measured viral suppression. The baseline-adjusted parameter (Eq2÷Eq3) can also be estimated with empirical proportions. Specifically, we would generate covariate strata-specific estimates by taking empirical means, and then combine by standardizing across strata.

A similar approach was used in the PopART Universal-Test-and-Treat trial with stratification factors including sex, age group and community.^17,33,34^ This approach corresponds to G-computation when fully-saturated regressions are used to estimate the conditional probability of the outcome, given measurement and the adjustment set (i.e. the “outcome regression”).^28,35,36^ It is further equivalent to inverse-weighting when fully-saturated regressions are used to estimate the conditional probability of measurement, given the adjustment set (i.e. the “propensity score”).^37–39^

When the adjustment set is higher dimensional, such as for our fully-adjusted parameter (Eq4÷Eq5), alternative approaches are needed to smooth over values of the covariates with weak support. We could, for example, use logistic regression with two-way interactions to estimate the propensity scores for inverse-weighting. This approach was used in a sensitivity analysis in the Ya Tsie Universal-Test-and-Treat trial.^18,40^

Another approach is targeted maximum likelihood estimation (TMLE), which offers efficiency gains over inverse-weighting and allows for flexible adjustment for a large set of covariates through machine learning.^24,41^ TMLE combines estimates of outcome regression with an estimate of the propensity score. (We refer the reader to ^42^ and ^43^ for an introduction.) TMLE is double robust - it is consistent if either the outcome regression is consistently estimated or the propensity score is consistently estimated. TMLE is also a substitution estimator, potentially improving robustness under strong confounding or rare outcomes.^44–47^

### Implementation

In SEARCH, the primary approach used TMLE to estimate the fully-adjusted parameter (Eq4÷Eq5). Within TMLE, Super Learner was implemented to estimate the outcome regressions and propensity scores.^48^ Super Learner is an ensemble, machine learning method using cross-validation to build the optimal combination of predictions from a library of candidate algorithms. We implemented TMLE fully stratified on community, allowing the outcome regressions and propensity scores to vary by community.

For comparison, we also present the use of empirical proportions to estimate the unadjusted parameter (Eq1), and the baseline-adjusted parameters (Eq2÷Eq3) controlling for sex, age group (15-19 years, 20-29 years, 30-39 years, 40-49 years, 50-59 years, and 60+ years), and community.

Statistical inference was obtained with influence curve standard errors, treating the community as the unit of independence. Analyses were conducted in R_v.3.6.1 with the ltmle_v1.1-0 and SuperLearner_v2.0-25 packages.^49–51^

## RESULTS

The baseline characteristics of the study participants have been described elsewhere.^8,9,25^ In brief, approximately one-third of the 79,818 residents enumerated in the baseline census were from each study region, and nearly half of participants were aged 15-30 years; men comprised 45% (Supplementary Table 1). HIV status was determined on 89% (71,402) of residents at baseline (Table 1). After baseline, knowledge of HIV serostatus remained high with 77% of residents (69,175/90,047) seen at population-level testing at Year 1, 75% (71,577/95,599) at Year 2, and 81% (80,390/99,186) at Year 3. There were no obvious demographic differences between the enumerated population and those with known HIV serostatus (Supplementary Table 1).

**Table 1:**
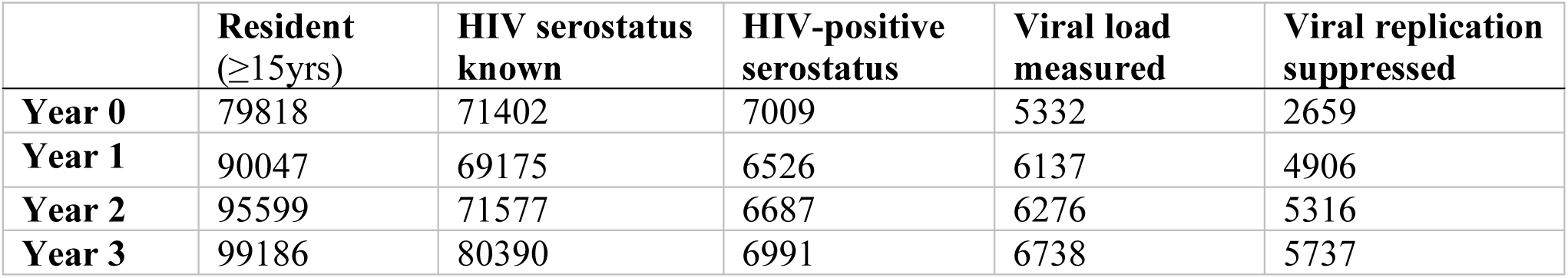
Number and coverage of residents contributing to unadjusted estimates of population-level HIV viral suppression at the time of annual testing. Each column is a subset of the former. Changes in annual population size are due to additions from in-migrants and aging-in, and due to subtractions from death and outmigration. Years refer to time since study baseline, which varied by community (Year 0 ranging from June 2013 to June 2014).

|  | **Resident**<br>**(≥15yrs)** | **HIV serostatus**<br>**known** | **HIV-positive**<br>**serostatus** | **Viral load**<br>**measured** | **Viral replication**<br>**suppressed** |
| --- | --- | --- | --- | --- | --- |
| **Year 0** | 79818 | 71402 | 7009 | 5332 | 2659 |
| **Year 1** | 90047 | 69175 | 6526 | 6137 | 4906 |
| **Year 2** | 95599 | 71577 | 6687 | 6276 | 5316 |
| **Year 3** | 99186 | 80390 | 6991 | 6738 | 5737 |

Viral loads were measured for 76% of baseline HIV-positive residents (Table 1). Missing viral loads were more common at baseline due to early assay failures.^26^ Despite ∼95% coverage of viral load measurement for the remaining years, baseline and time-varying characteristics differed for HIV-positive persons with measured versus missed HIV RNA levels (Table 2). In particular, HIV-positive women were more likely to have their viral load measured than HIV-positive men. After baseline, adolescents (15-24years) were more likely to be missed than older adults (25+years). Viral load measurement also differed notably by the time-varying characteristics; HIV-positive persons who were previously aware of their status, had evidence of starting ART, or had a history of suppressing viral replication were more likely to have their viral load measured than their counterparts.

**Table 2:**
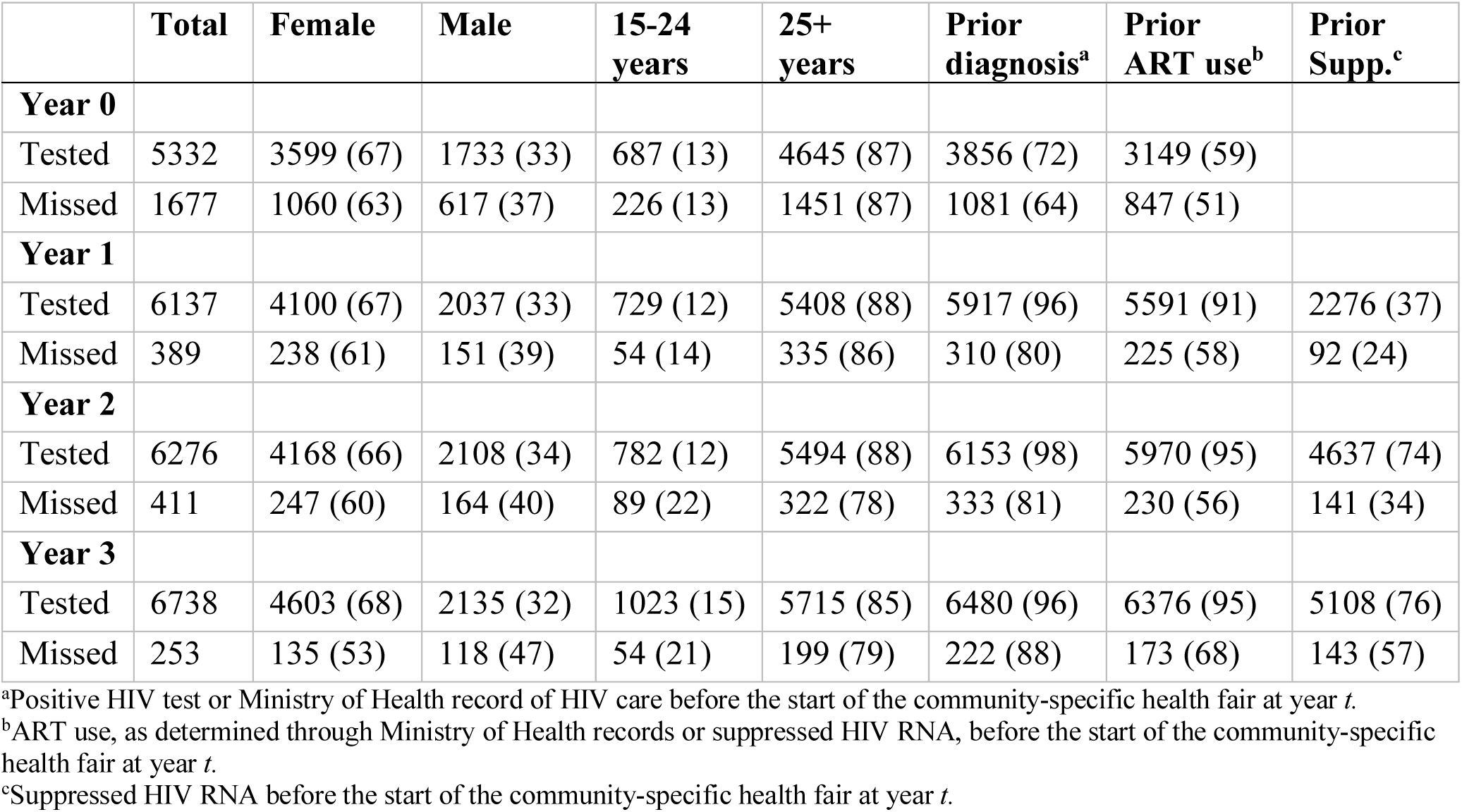
Select baseline and time-varying characteristics of HIV-positive residents by year and by viral load measurement. Metrics in N (%).

|  | **Total** | **Female** | **Male** | **15-24**<br>**years** | **25+**<br>**years** | **Prior**<br>**diagnosis<sup>a</sup>** | **Prior**<br>**ART use<sup>b</sup>** | **Prior**<br>**Supp.<sup>c</sup>** |
| --- | --- | --- | --- | --- | --- | --- | --- | --- |
| **Year 0** |  |  |  |  |  |  |  |  |
| Tested | 5332 | 3599 (67) | 1733 (33) | 687 (13) | 4645 (87) | 3856 (72) | 3149 (59) |  |
| Missed | 1677 | 1060 (63) | 617 (37) | 226 (13) | 1451 (87) | 1081 (64) | 847 (51) |  |
| **Year 1** |  |  |  |  |  |  |  |  |
| Tested | 6137 | 4100 (67) | 2037 (33) | 729 (12) | 5408 (88) | 5917 (96) | 5591 (91) | 2276 (37) |
| Missed | 389 | 238 (61) | 151 (39) | 54 (14) | 335 (86) | 310 (80) | 225 (58) | 92 (24) |
| **Year 2** |  |  |  |  |  |  |  |  |
| Tested | 6276 | 4168 (66) | 2108 (34) | 782 (12) | 5494 (88) | 6153 (98) | 5970 (95) | 4637 (74) |
| Missed | 411 | 247 (60) | 164 (40) | 89 (22) | 322 (78) | 333 (81) | 230 (56) | 141 (34) |
| **Year 3** |  |  |  |  |  |  |  |  |
| Tested | 6738 | 4603 (68) | 2135 (32) | 1023 (15) | 5715 (85) | 6480 (96) | 6376 (95) | 5108 (76) |
| Missed | 253 | 135 (53) | 118 (47) | 54 (21) | 199 (79) | 222 (88) | 173 (68) | 143 (57) |

<sup>a</sup>Positive HIV test or Ministry of Health record of HIV care before the start of the community-specific health fair at year *t*.
<sup>b</sup>ART use, as determined through Ministry of Health records or suppressed HIV RNA, before the start of the community-specific health fair at year *t*.
<sup>c</sup>Suppressed HIV RNA before the start of the community-specific health fair at year *t*.

Estimates of population-level suppression did vary meaningfully by identifiability assumption (Figure 2). At baseline, the unadjusted approach suggested that half of all HIV-positive residents had suppressed viral replication (50%; 95%CI: 46-54%). Stratifying on age group, sex, and community slightly reduced the estimate to 49% (95%CI: 45-54%), and the most conservative estimate of 42% (95%CI: 38-46%) was obtained after adjusting for the full set of baseline and time-varying characteristics.

**Figure 2:**
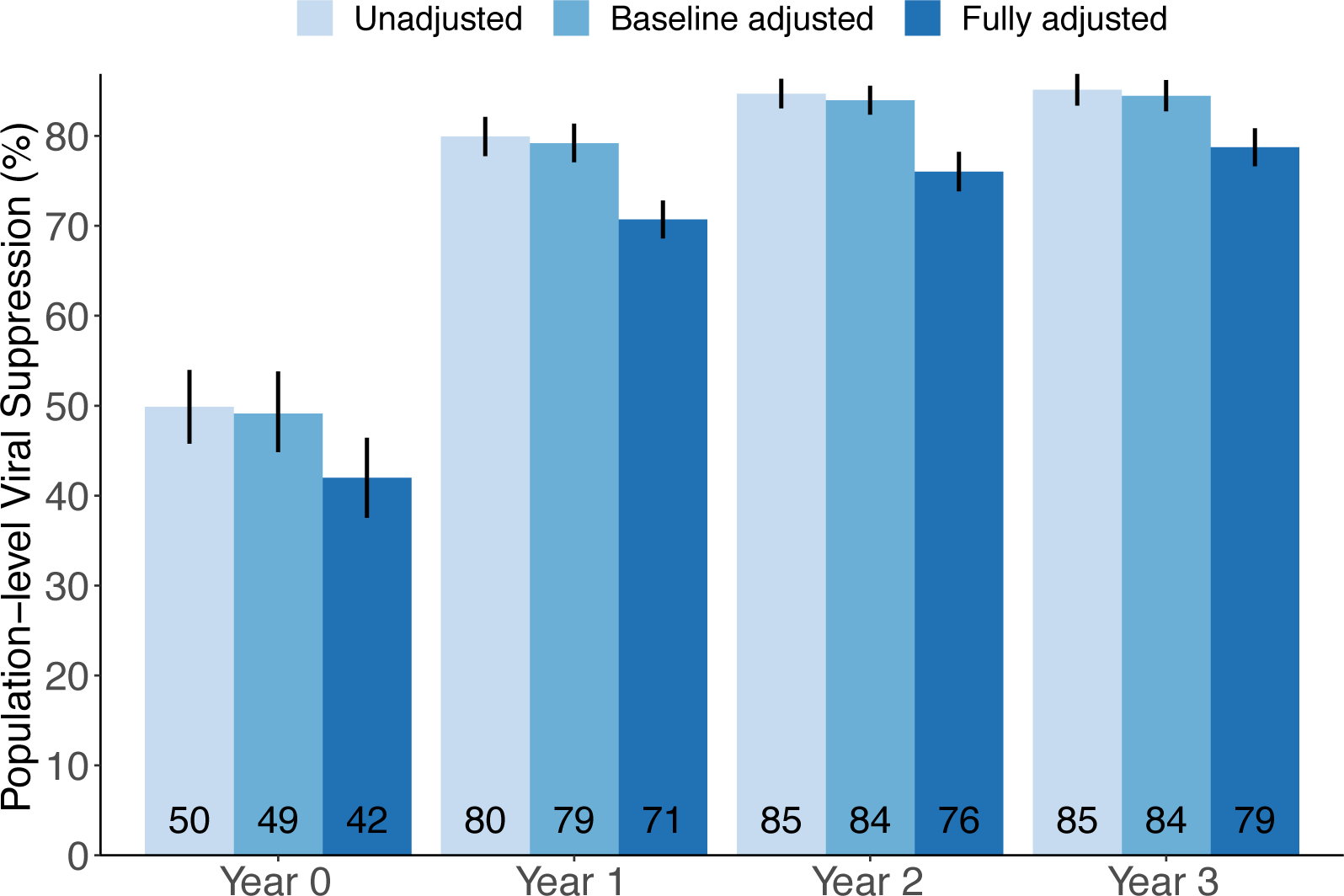
Estimates of population-level HIV viral suppression at the time of annual testing in the intervention arm of the SEARCH trial. Estimates were obtained with the empirical mean among those measured (“Unadjusted”), stratifying on sex, age group and community (“Baseline adjusted”), and using targeted maximum likelihood estimation (TMLE) with Super Learner to adjust for both baseline and time-varying characteristics (“Fully adjusted”). Black vertical bars indicate 95% confidence intervals.

Deviations between estimates of population-level suppression were pronounced in subsequent years (Figure 2). The unadjusted approach suggested that 80% (95%CI: 78-82%) of all HIV-positive residents were suppressed at Year 1, 85% (95%CI: 83-86%) at Year 2, and 85% (95%CI: 83-87%) at Year 3. Estimates adjusted for baseline covariate-strata were similar: 79% (95%CI: 77-81%) at Year 1, 84% (95%CI: 82-86%) at Year 2, and 84% (95%CI: 83-86%) at Year 3. Fully adjusted estimates were the most conservative: 71% (95%CI: 69-73%) at Year 1, 76% (95%CI: 74-78%) at Year 2, and 79% (95%CI: 77-81%) at Year 3.

## DISCUSSION

In an open cohort of nearly 100,000 residents in rural Kenya and Uganda, we compared three approaches for estimating population-level HIV viral suppression: (i) an unadjusted approach, the empirical proportion among those measured; (ii) stratification on age group, sex, and community; and (iii) TMLE with Super Learner to adjust for the full set of baseline and time-varying covariates. Despite high coverage of out-of-facility testing, estimates diverged by identifiability assumptions. The unadjusted approach consistently yielded the highest estimates; the fully-adjusted approach consistently yielded the lowest estimates.

In the SEARCH study, HIV serostatus and HIV RNA viral levels were obtained through multidisease testing at health fairs with follow-up for non-participants.^25^ Unlike clinic-based ascertainment, this approach reaches HIV-positive persons who are in-care as well as newly diagnosed and previously diagnosed but out-of-care.^9,16–18^ As a result, the MCAR assumption may seem reasonable.^1–4^ However, deviations between the unadjusted estimates and adjusted ones suggest there were meaningful differences in the population measured and population missed with respect to, among other factors, prior diagnosis, ART use, and viral suppression.

Both adjusted approaches were built on the missing-at-random (MAR) assumption: knowledge of HIV status and viral load measurement were only a function of observed characteristics.^1–4^ When controlling for baseline covariates, we fully stratified on sex, age group, and community; as a result, this was equivalent to a fully non-parametric approach for the outcome regression in G-computation and to a fully non-parametric approach for the propensity score in inverse-weighting. Beyond age, sex, and community, there were, however, additional differences between those with measured versus missing viral loads, including differences in post-baseline variables; specifically, persons without prior diagnosis, ART initiation, or viral suppression were less likely to have their viral load measured.

Therefore, our primary approach in SEARCH was to weaken the identifiability assumptions by adjusting for a larger set of baseline and time-updated covariates. TMLE with Super Learner was used to flexibly estimate the conditional probability of HIV seropositivity, conditional probability of suppression, and conditional probabilities of measurement.^8,9,27^

In this example, the identification choice has implications for policy-making and targeting resources. Both the unadjusted and baseline-adjusted approaches suggested the UNAIDS 90-90-90 target (73%-suppression) was surpassed within one year of the intervention and the UNAIDS 95-95-95 target (86%-suppression) was nearly achieved by the trial’s close.^5^ In contrast, the estimates controlling for time-updated covariates indicated the 90-90-90 target was achieved after two years, but there still was a substantial gap to the 95-95-95 target.

In summary, estimates of population-level HIV viral suppression continue to be the benchmark in assessing programmatic success in epidemic control. In four cross-sectional analyses of 79,818-99,186 participants in the intervention arm of SEARCH, we demonstrated the impact of assumptions on incomplete measurement that can occur even in “Big Data” settings. We recommend adjustment for a large set of baseline and time-varying covariates that potentially influence both measurement and underlying status; TMLE with Super Learner is one approach to performing such adjustments.

### Appendix: UNAIDS 90-90-90 target and population-level suppression

For the moment assume complete measurement, and let *HIV*_*t*_ be an indicator of HIV-positive serostatus at time *t*; *Dx*_*t*_ be an indicator of having an HIV diagnosis by time *t; ART*_*t*_ be an indicator of antiretroviral therapy (ART) use at time *t*, and *Supp*_*t*_ be an indicator of suppressed viral replication at time *t*. The UNAIDS 90-90-90 targets are a series of proportions or conditional probabilities:^5^

% of all HIV-positives who are diagnosed (first-90):

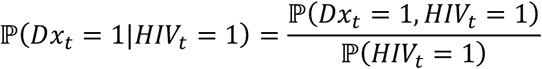

% of diagnosed who are on ART (second-90):

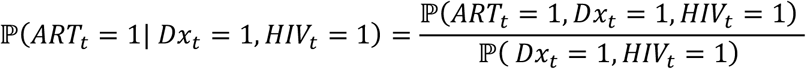

% on ART who are currently suppressed (third-90):

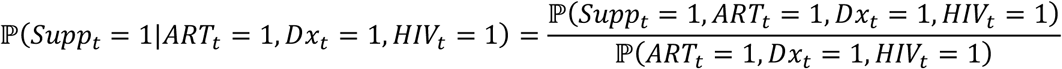

Multiplying together the three “90s” yields the proportion of all HIV-positive persons who are currently suppressed (i.e. population-level suppression):

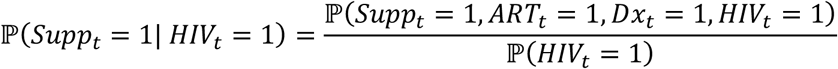

Since each numerator and denominator is a population-level proportion, we can equivalently express the targets as follows: first-90=(number previously diagnosed)/(number HIV-positive), second-90=(number on ART)/(number previously diagnosed), third-90=(number virally suppressed)/(number on ART), and population-level suppression=(number virally suppressed)/(number HIV-positive).

Therefore, one could directly estimate population-level suppression, as we demonstrated here, or instead estimate each 90-90-90 target and multiply. These two approaches should yield identical results, as demonstrated in our previous work.^8,9,27^ However, deviations between the direct estimate and the multiplied-one can occur when making the missing-completely-at-random (MCAR) assumption.^1–4^ Specifically, under MCAR, the denominators of the third-90 and population-level suppression become conditional on having a viral load measured, which is almost always a subset of the population on ART and a subset of the population who is HIV-positive.

## Data Availability

Data sufficient to reproduce the study findings will be made available approximately one year after completion of the ongoing trial (NCT01864603). Further inquiries can be directed to the SEARCH Scientific Committee at.

## Acknowledgements

We thank the Ministry of Health of Uganda and of Kenya; our research teams and administrative teams in San Francisco, Uganda, and Kenya; collaborators and advisory boards; and especially all the communities and participants involved in the study.

## Notes

Conflicts of Interest: There is no conflict of interest

Sources of Funding: This work was supported by grant numbers U01AI099959, UM1AI068636, and R01AI074345-06A1 from National Institute of Allergy and Infectious Diseases at the National Institutes of Health; by the President’s Emergency Plan for AIDS Relief; and by Gilead Sciences, which provided Truvada®.

### Competing Interest Statement

The authors have declared no competing interest.

### Clinical Trial

NCT01864603

### Funding Statement

This work was supported by grant numbers U01AI099959, UM1AI068636, and R01AI074345-06A1 from National Institute of Allergy and Infectious Diseases at the National Institutes of Health; by the President’s Emergency Plan for AIDS Relief; and by Gilead Sciences, which provided Truvada®.

